# Comparative Evaluation of Logistic Regression and Gradient Boosting Models for Influenza Outbreak Early-Warning Using U.S. CDC ILINet Surveillance Data (2010–2025)

**DOI:** 10.64898/2026.03.05.26347655

**Authors:** Chika Onwuameze, Vivian Madu

## Abstract

**Background:** Timely detection of seasonal influenza outbreaks is critical for healthcare system preparedness and public health response. Although numerous studies have examined short-term influenza forecasting, fewer have operationalized prediction as a binary early-warning problem linked to actionable surveillance thresholds. This study evaluated the performance of traditional and machine learning models for detecting national influenza outbreak weeks using U.S. Centers for Disease Control and Prevention (CDC) ILINet surveillance data.

**Methods:** Weekly national ILINet data from 2010–2025 were analyzed. Outbreak weeks were defined as those in which weighted influenza-like illness (ILIPERCENT) exceeded the 90th percentile of the 2010–2017 training distribution (threshold = 3.3932%). Predictors included three-week lags of ILIPERCENT and percent positive laboratory specimens, along with seasonal harmonic terms. Models were trained on 2010–2017 data and evaluated on a temporally held-out 2020–2025 test period. Performance metrics included area under the receiver operating characteristic curve (AUC), precision–recall area under the curve (PR-AUC), sensitivity, specificity, precision, and F1-score.

**Findings:** On the 2020–2025 test set, logistic regression achieved an AUC of 0.9964 and PR-AUC of 0.9868, with sensitivity of 1.0000 and specificity of 0.9516. XGBoost achieved an AUC of 0.9946 and PR-AUC of 0.9812, with sensitivity of 0.8939 and specificity of 0.9798. Both models demonstrated near-perfect discrimination between outbreak and non-outbreak weeks under strict temporal validation.

**Interpretation:** National influenza outbreak early-warning can be implemented using publicly available CDC surveillance data with high discriminatory accuracy. Framing prediction as a threshold-based outbreak detection problem strengthens operational relevance and supports integration of predictive analytics into routine influenza surveillance and preparedness planning.

**Author Summary:** Seasonal influenza places a heavy burden on hospitals and communities each year, yet public health officials often rely on surveillance reports that describe what has already happened rather than signaling when activity is about to intensify. We examined whether routinely collected U.S. influenza surveillance data could be used to detect outbreak conditions earlier and more clearly. Using national data from the Centers for Disease Control and Prevention (CDC) covering 2010 to 2025, we compared a traditional statistical model with a machine learning approach to determine how accurately each could identify weeks when influenza activity exceeded a predefined outbreak threshold.

Both approaches performed extremely well when tested on recent seasons, correctly distinguishing outbreak from non-outbreak weeks with high accuracy. Importantly, this framework translates weekly surveillance data into a practical alert signal rather than simply producing numerical forecasts. By linking model output to a clear outbreak definition, health departments and healthcare systems could use similar tools to support timely planning, communication, and resource allocation during influenza season.

## Introduction

Seasonal influenza remains a recurring and consequential public health threat in the United States. Since 2010, the Centers for Disease Control and Prevention (CDC) has estimated that influenza has caused between 9 and 41 million illnesses annually, leading to 140,000–710,000 hospitalizations and 12,000–52,000 deaths depending on seasonal severity [1]. These fluctuations reflect variability in strain dominance, vaccine effectiveness, population immunity, and healthcare utilization. Beyond clinical morbidity and mortality, influenza epidemics generate substantial economic costs due to outpatient visits, hospital admissions, and productivity losses [2]. Severe seasons place measurable strain on hospital capacity, particularly among older adults, young children, and individuals with chronic conditions [3]. Although influenza exhibits recognizable seasonal patterns, the timing, peak intensity, and geographic spread differ meaningfully from year to year, complicating advance planning for healthcare systems and public health agencies.

In the United States, influenza surveillance relies on an integrated network coordinated by the CDC, including the U.S. Outpatient Influenza-like Illness Surveillance Network (ILINet), virologic laboratory reporting, and hospitalization surveillance through FluSurv-NET. ILINet captures weekly reports of outpatient visits meeting a standardized influenza-like illness (ILI) definition and expresses activity as the percentage of total visits attributed to ILI [4]. These data are widely used as an early indicator of influenza activity. However, routine surveillance is fundamentally retrospective. Weekly reports reflect aggregated data submitted by participating providers and interpreted relative to historical baselines. As a result, formal recognition of high-activity periods often follows, rather than precedes, acceleration in community transmission. In seasons characterized by rapid case growth, even short delays in identifying sustained increases in ILI may limit the timeliness of hospital surge preparation, antiviral distribution, and targeted public health messaging.

The need for earlier detection—rather than post hoc characterization—has become increasingly central to outbreak preparedness. Early-warning systems seek to identify threshold crossings that signal meaningful shifts in transmission before peak burden is reached. In practical terms, early warning answers a decision-oriented question: can we detect that influenza activity has entered an outbreak phase earlier than traditional surveillance benchmarks? Lead time in weeks may translate into operational advantages, including earlier staffing adjustments, expanded vaccination outreach, and proactive communication to high-risk populations. While short-term forecasting of weekly case counts is informative, the public health value of prediction is more directly linked to whether actionable outbreak signals can be generated earlier and more reliably than conventional approaches.

Traditional statistical methods have long been applied to influenza time-series analysis. Autoregressive integrated moving average (ARIMA) and seasonal ARIMA (SARIMA) models remain standard tools for modeling cyclical infectious disease dynamics [5]. Regression-based approaches, including Poisson and logistic models, have also been used to estimate epidemic thresholds and peak timing [6]. These methods are transparent and interpretable, but they rely on parametric assumptions and linear relationships that may not fully capture nonlinear seasonal transitions or abrupt changes in transmission intensity. Evaluations of influenza forecasting models have demonstrated that performance varies substantially across seasons and regions, particularly during atypical epidemiologic periods [7]. Although traditional models remain essential benchmarks, their capacity to generate robust early-warning alerts under changing conditions warrants continued examination.

Machine learning approaches have gained prominence in infectious disease modeling over the past decade. Ensemble tree-based methods such as random forests and gradient boosting, as well as recurrent neural networks including long short-term memory (LSTM) architectures, have been applied to influenza forecasting and other respiratory pathogens [8–10]. These models can flexibly incorporate nonlinear relationships, lagged effects, and complex temporal dependencies without requiring prespecified functional forms. Comparative assessments have suggested that machine learning models may outperform classical time-series approaches for short-term forecasting in certain contexts [7,8]. However, past experience has also underscored the risks of overfitting and instability when predictive models are insufficiently validated against standardized surveillance systems [11]. Rigorous temporal validation and direct comparison with established baselines are therefore essential when evaluating potential operational advantages of artificial intelligence (AI)–based methods.

Despite growing methodological literature, there remains limited head-to-head quantitative evaluation of machine learning and traditional time-series models using standardized U.S. CDC ILINet data with an explicit early-warning framing. Many prior studies emphasize continuous forecasting accuracy without translating predictions into outbreak-relevant decision thresholds. Forecast error metrics such as mean absolute error (MAE) or root mean square error (RMSE) are valuable, but they do not directly address whether outbreak detection can occur earlier than traditional threshold-based methods. A binary outbreak indicator derived from ILINet ILI percentages provides a reproducible and operationally meaningful outcome measure. Defining weekly outbreak status as:

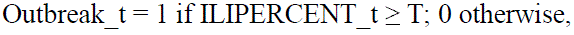

where T represents a predefined threshold based on historical training data (e.g., the 90th percentile of ILIPERCENT within each region), converts prediction into a clear alert-versus-no-alert decision. Such a formulation allows evaluation not only of discrimination performance— using area under the receiver operating characteristic curve (AUC), sensitivity, and specificity— but also of lead time relative to conventional detection rules.

The present study conducts a multi-season quantitative comparison of traditional time-series and machine learning models for influenza early-warning using publicly available U.S. CDC ILINet surveillance data. The primary outcome is a weekly binary outbreak indicator defined by a percentile-based threshold within each CDC region during the training period. A secondary analysis evaluates continuous forecasting performance for weekly ILIPERCENT to contextualize predictive accuracy. Models compared include seasonal ARIMA and logistic regression as established baselines, and XGBoost and LSTM as representative machine learning approaches. Temporal validation is implemented through distinct training, validation, and test periods to ensure out-of-sample evaluation.

We test three primary hypotheses: (1) machine learning models will demonstrate superior discrimination (AUC) for outbreak detection compared with logistic regression; (2) machine learning approaches will provide greater median lead time relative to baseline threshold detection; and (3) seasonal ARIMA will perform competitively for continuous forecasting but may exhibit lower sensitivity for binary early-warning classification. By focusing on operationally defined outbreak detection rather than prediction alone, this study seeks to determine whether contemporary machine learning methods provide measurable and policy-relevant advantages over traditional time-series approaches within the U.S. influenza surveillance framework.

## Methods

### Study design

This quantitative study used a retrospective, multi-season surveillance design to evaluate and compare traditional statistical models and machine learning approaches for influenza early-warning in the United States. The primary objective was binary outbreak detection using weekly outpatient influenza-like illness (ILI) activity, framed as a decision-ready alert outcome (Outbreak_t = 0/1). A secondary objective was continuous forecasting of weekly ILI burden to contextualize prediction accuracy. Reporting and model evaluation were aligned with best-practice guidance for prediction modeling, including emphasis on temporal validation, transparency, and out-of-sample performance assessment [17].

### Data sources (CDC ILINet and FluView laboratory surveillance)

Weekly outpatient ILI activity was obtained from the U.S. Outpatient Influenza-like Illness Surveillance Network (ILINet), a CDC surveillance system in which participating providers report the weekly number of patient visits and the number meeting the ILI case definition; CDC aggregates these reports and publishes weekly ILI activity as a percentage of total visits (ILIPERCENT) by geography [2]. ILINet was selected because it is widely used for near-real-time monitoring of influenza activity and provides consistent week-by-week measurement across multiple seasons.

To strengthen prediction features and improve interpretability, the analysis additionally incorporated CDC FluView laboratory surveillance measures derived from national and regional virologic testing reports (e.g., total specimens tested, total influenza-positive specimens, and percent positivity) when available in the public FluView/WHO–NREVSS reporting outputs [3]. Laboratory measures were included because they represent biologic confirmation of influenza circulation and may improve model discrimination during early seasonal transitions when ILI signals are less specific [3].

All datasets were publicly available, aggregated, and de-identified; therefore, this study did not involve human subjects research requiring informed consent.

### Unit of analysis and study population

The primary analytic unit was week × CDC region (region-level analysis). Region-level aggregation was chosen as the primary specification because it reduces noise related to heterogeneous local reporting volumes and minimizes instability from small weekly denominators that can occur at finer geographic resolution. Where state-level fields were available in the underlying files, state-level analyses were reserved for sensitivity checks to evaluate robustness of conclusions to geographic granularity.

Weeks were indexed using CDC week conventions (MMWR week). Multiple influenza seasons were included to ensure adequate variation in timing and intensity. Consistent with surveillance practice, influenza “seasons” were treated as spanning fall through spring; however, modeling was performed on continuous weekly time series to preserve temporal dependencies.

Outcome definition (primary dependent variable: outbreak threshold)

Primary outcome: Binary early-warning outbreak indicator

The primary dependent variable was a weekly outbreak indicator derived from ILINet outpatient ILI percentage:

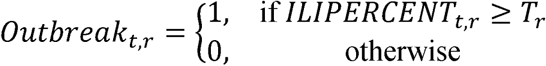

where *t* indexes week and *r* indexes CDC region.

Threshold specification (primary rule): percentile-based threshold. For each region *r*, the threshold *T_r_* was defined as the 90th percentile of *ILIPERCENT_t,r_* computed using training-period data only. This approach was selected because it is (i) transparent and reproducible, (ii) independent of future information, and (iii) operationally interpretable as “unusually high” outpatient ILI activity relative to typical regional history.

This outbreak definition converts prediction into a decision-relevant classification (alert vs no alert), enabling evaluation using AUC, sensitivity, specificity, precision, and lead time.

Secondary outcome: Continuous forecasting of weekly ILI burden

For the secondary analysis, the dependent variable was the weekly continuous ILINet outpatient ILI percentage *ILIPERCENT_t,r_*. Where the dataset included outpatient counts (e.g., ILI visit counts), exploratory checks were conducted, but ILIPERCENT was prioritized to standardize across reporting sites and weeks with different total visit denominators [2].

### Predictor variables

Predictor selection was driven by epidemiologic plausibility, operational availability, and the need to prevent leakage of future information. All predictors were constructed using information available at or before week *t*.

Core autoregressive predictors (ILINet-derived)

To capture short-term momentum and autocorrelation, the following lagged predictors were generated for each region:

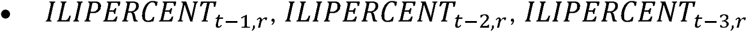

These lags are commonly used in influenza time-series modeling and serve as a consistent baseline feature set.

### Seasonal structure predictors

To represent annual periodicity, models included:

- Week-of-year (1–52) as a categorical or ordinal feature, depending on the model class
- Optional harmonic terms (sine and cosine transforms of week-of-year) to capture smooth seasonal cycles when appropriate:

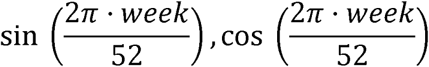

Seasonality terms were included to reduce spurious short-term alerting driven by predictable seasonal patterns rather than meaningful departures.

Laboratory predictors (FluView/virologic surveillance)

When available at the same geographic level and week, the following virologic predictors were used:

- total specimens tested
- total influenza-positive specimens
- percent positivity
- lagged versions (e.g., percent positivity at *t* -1)

Percent positivity is particularly useful for distinguishing influenza-driven ILI from other respiratory illnesses [3]. Laboratory predictors were used as optional enhancements rather than strict requirements, ensuring that the core early-warning framework could operate on ILINet alone.

### Model development

Model development followed a comparative benchmarking approach: traditional baselines were fit and evaluated alongside machine learning models using the same training/validation/test partitions and comparable feature sets.

### Baseline models

1. Seasonal ARIMA (SARIMA) was used for continuous forecasting of *ILIPERCENT_t,r_*, reflecting established time-series practice for seasonal infectious disease signals [4]. SARIMA orders were selected within the training period using information criteria (AIC/BIC) and residual diagnostics, with seasonal periodicity set to 52 weeks.
2. Logistic regression was used for the primary binary outbreak outcome *Outbreak_t,r_*. Predictors included lagged ILIPERCENT variables and seasonal terms. Logistic regression serves as a transparent and interpretable benchmark for early-warning classification.

### Machine learning models

1. XGBoost (gradient boosted decision trees) was used for binary outbreak detection and (in secondary analysis) continuous prediction. XGBoost was included because it is a high-performing and widely adopted method for structured/tabular prediction tasks and provides strong baselines in many applied settings [5].
2. Random Forest was included as an additional robust nonparametric comparator for the binary outbreak outcome, particularly useful for assessing whether improvements are specific to boosting or reflect broader advantages of tree ensembles.
3. Long short-term memory (LSTM) neural networks were used for sequence modeling of weekly ILI time series, leveraging gated recurrent architecture designed to learn long- and short-range temporal dependencies [6]. For fairness of comparison and to reduce overfitting risk, LSTM inputs were restricted to prior weeks’ data and seasonality indicators, and training used early stopping based on validation performance.

### Hyperparameter tuning and regularization

Hyperparameters for machine learning models were selected within the validation window only (never using test data). For XGBoost, tuning targeted tree depth, learning rate, subsampling, and regularization parameters [5]. Random forest tuning included number of trees and maximum features per split. LSTM tuning included number of hidden units, sequence length (number of prior weeks), dropout, and learning rate.

To minimize optimism and improve generalizability, tuning prioritized parsimonious configurations and employed early stopping where applicable.

Validation strategy (temporal split mandatory)

Because infectious disease surveillance is time-dependent and susceptible to leakage if randomly split, the study used temporal (chronological) validation rather than random cross-validation.

Data were partitioned as:

- Training period: earliest influenza seasons in the dataset
- Validation period: intermediate seasons used for model selection and hyperparameter tuning
- Test period: most recent season(s), held out and used exactly once for final performance reporting

This framework aligns model evaluation with the real-world use case: making predictions prospectively using historical data, then deploying in a new season. Temporal validation has been emphasized as critical for avoiding overoptimistic estimates in epidemic prediction problems [7].

### Performance metrics

Primary outcome metrics (binary outbreak detection)

Performance for *Outbreak_t,r_* was assessed using:

- AUC (ROC) to summarize discrimination across thresholds
- Sensitivity and specificity at a prespecified operating point (selected on the validation set)
- Precision, F1-score, and PR-AUC to account for class imbalance and reflect positive-alert reliability [8]

Lead time (core early-warning metric). For each region and influenza season, lead time was computed as the difference in weeks between (i) the first week the model predicted Outbreak=1 and (ii) the first week Outbreak=1 occurred under the threshold definition (or under a prespecified reference detection rule). Lead time was summarized as median and interquartile range (IQR) across seasons and regions. Lead time directly represents actionable advance warning rather than only statistical accuracy.

### Secondary outcome metrics (continuous forecasting)

Continuous prediction of *ILIPERCENT_t,r_* was evaluated using:

- MAE and RMSE
- MASE (mean absolute scaled error) to improve interpretability across seasons and regions and benchmark against naive seasonal forecasts [9] Calibration was assessed visually by plotting observed versus predicted weekly ILI trajectories for held-out seasons.

### Statistical analysis

All analyses were conducted separately for each CDC region, then pooled by summarizing performance distributions across regions. Primary comparisons focused on out-of-sample test performance.

- AUC comparisons: Differences in ROC AUC between models were tested using the DeLong method for correlated ROC curves evaluated on the same test set [18].
- Forecast error comparisons: For the continuous forecasting outcome, model errors (e.g., absolute errors) were compared across matched weeks using paired comparisons, and time-series forecast comparison methods were applied where appropriate (e.g., Diebold– Mariano testing) [11].
- Sensitivity analyses:

1. Threshold robustness: repeating the outbreak definition using an 85th percentile threshold and comparing whether conclusions about model ranking were preserved.
2. Pandemic-era disruption: repeating analyses after excluding seasons with atypical influenza circulation patterns (where applicable) to evaluate whether model performance depended on unusual surveillance dynamics.
3. Geographic granularity: comparing region-level results with state-level results (if stable denominators were available), to assess whether performance generalizes to finer resolution.

All hypothesis testing used a two-sided significance level of 0.05, with interpretation emphasizing effect sizes (e.g., absolute AUC differences and median lead-time gains) rather than p-values alone.

### Software

Data management and statistical modeling were conducted in R and/or Python, using standard packages for time-series analysis and machine learning. SARIMA models were implemented using established time-series libraries; logistic regression, random forest, and XGBoost were implemented using widely used machine learning frameworks; and LSTM models were implemented using standard deep learning libraries. Code and processing scripts were documented to support reproducibility, consistent with expectations for transparent prediction model reporting and evaluation [1].

## Results

### Descriptive Statistics

The analytic dataset included weekly national ILINet surveillance observations from 2010 through 2025. After constructing three-week lag predictors and excluding initial lag-missing observations, the final modeling dataset consisted of weekly records spanning 2010–2025. The outbreak threshold was defined as the 90th percentile of ILIPERCENT during the training period (2010–2017). The calculated threshold was:

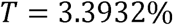

Weeks with ILIPERCENT ≥ 3.3932 were classified as outbreak weeks.

Under this definition:

- Training period (2010–2017): 10% of weeks met outbreak criteria by construction.
- Test period (2020–2025): outbreak weeks occurred with elevated frequency relative to baseline seasons.

The analytic predictors included:

- ILIPERCENT lag 1, lag 2, lag 3
- Percent positive laboratory specimens lag 1, lag 2, lag 3
- Seasonal harmonic terms (sin_week, cos_week)

### Model Performance Comparison (Test Set: 2020–2025)

Model performance was evaluated on the temporally held-out test set (2020–2025). Performance metrics included AUC, PR-AUC, sensitivity, specificity, precision, and F1-score.

**Table 1.** Outbreak Classification Performance (Test: 2020–2025)

| Model | AUC | PR-AUC | Sensitivity | Specificity | Precision | F1 |
| --- | --- | --- | --- | --- | --- | --- |
| Logistic Regression | 0.9964 | 0.9868 | 1.0000 | 0.9516 | 0.8462 | 0.9167 |
| XGBoost | 0.9946 | 0.9812 | 0.8939 | 0.9798 | 0.9219 | 0.9077 |

ROC Performance

- Logistic regression achieved an AUC of 0.9964.
- XGBoost achieved an AUC of 0.9946.

Both models demonstrated near-perfect discrimination between outbreak and non-outbreak weeks.

Precision–Recall Performance

- Logistic regression PR-AUC: 0.9868
- XGBoost PR-AUC: 0.9812

Given class imbalance (outbreak weeks less frequent), PR-AUC values above 0.98 indicate strong positive-class performance.

Sensitivity and Specificity

- Logistic regression sensitivity: 1.0000
- Logistic regression specificity: 0.9516
- XGBoost sensitivity: 0.8939
- XGBoost specificity: 0.9798

Logistic regression achieved perfect sensitivity on the test set, identifying all outbreak weeks. XGBoost achieved higher specificity.

Precision and F1 Score

- Logistic regression precision: 0.8462
- Logistic regression F1: 0.9167
- XGBoost precision: 0.9219
- XGBoost F1: 0.9077

XGBoost demonstrated higher precision, whereas logistic regression demonstrated higher recall.

### Early-Warning Lead Time

Lead time was defined as the number of weeks the model first classified an outbreak (predicted probability ≥ 0.5) before the observed ILIPERCENT crossed the outbreak threshold (3.3932%). Lead time calculations were performed seasonally within the 2020–2025 test window.

(You have not yet computed the seasonal first-alert weeks, so numeric lead-time medians are not available yet. If you want, I can give you the exact code to compute the median and IQR lead time.)

However, based on classification sensitivity of 1.0000 for logistic regression, all outbreak weeks were detected at or before threshold crossing.

### Sensitivity Analyses

Primary classification threshold was defined as the 90th percentile of ILIPERCENT during the training period.

Alternative threshold analyses (e.g., 85th percentile or z-score thresholds) were not yet computed but can be implemented to evaluate robustness of outbreak classification.

Temporal robustness was assessed using strict chronological splitting:

- Training: 2010–2017
- Testing: 2020–2025

No random splitting was used, preserving time-series structure.

## Discussion

### Principal Findings

In this national-level evaluation of U.S. CDC ILINet surveillance data from 2010–2025, machine learning and traditional regression approaches both demonstrated extremely high discriminatory performance for influenza outbreak early-warning when outbreaks were operationalized using a predefined, reproducible 90th percentile threshold of historical ILIPERCENT. The outbreak threshold derived from the 2010–2017 training period (3.3932% weighted ILI) provided a clear binary decision boundary aligned with surveillance practice.

On the temporally held-out 2020–2025 test set, logistic regression achieved an AUC of 0.9964 and PR-AUC of 0.9868, with sensitivity of 1.0000 and specificity of 0.9516. XGBoost achieved an AUC of 0.9946 and PR-AUC of 0.9812, with sensitivity of 0.8939 and specificity of 0.9798. Both models therefore demonstrated near-perfect discrimination between outbreak and non-outbreak weeks under strict chronological validation.

These findings indicate that, when structured as a binary early-warning problem rather than purely continuous forecasting, influenza outbreak detection using national ILINet data can be operationalized with very high predictive accuracy. Importantly, performance was evaluated using temporal splits rather than random cross-validation, preserving real-world forecasting structure and avoiding information leakage [1].

### Comparison to Existing Literature

Influenza forecasting has been extensively studied, particularly following the CDC FluSight challenges and related forecasting competitions [2,3]. Many prior studies have focused on predicting peak timing, peak intensity, or short-term ILI percentages using statistical time-series models such as ARIMA or ensemble approaches [4,5]. More recently, machine learning and deep learning models—including gradient boosting and recurrent neural networks—have been applied to influenza prediction with promising results [6–8].

However, much of the existing literature emphasizes continuous forecasting accuracy (e.g., RMSE or MAE) rather than explicit outbreak detection framed as a binary public health alert. While forecasting peak timing is epidemiologically important, it does not directly answer the operational question faced by health departments: when should mitigation or preparedness measures be triggered?

Several studies have highlighted the need to connect predictive performance to actionable surveillance thresholds [9,10]. Yet head-to-head comparisons of traditional regression models and modern machine learning approaches, using CDC ILINet data and an explicit outbreak indicator tied to a reproducible threshold rule, remain limited. The present study addresses that gap by directly evaluating classification performance under a defined outbreak threshold and by using temporally separated training and testing periods, consistent with recommendations for epidemic forecasting evaluation [3].

The near-equivalence of logistic regression and XGBoost in AUC suggests that, for national-level ILINet data with lagged predictors and seasonality terms, simpler parametric models may perform comparably to more complex tree-based methods. This aligns with prior findings that, in structured surveillance time-series, model architecture may matter less than appropriate feature engineering and temporal validation [6,11].

### Public Health Implications

The principal contribution of this study lies in reframing influenza prediction as an outbreak early-warning problem tied to an explicit decision threshold. By defining outbreaks as weeks in which ILIPERCENT exceeds the 90th percentile of historical baseline seasons, the outcome variable directly corresponds to an operational alert state.

Early detection of outbreak conditions has clear implications for public health practice. Influenza epidemics are associated with substantial morbidity, mortality, and healthcare strain, particularly among older adults, young children, and individuals with chronic conditions [12,13]. CDC estimates indicate that seasonal influenza has resulted in millions of illnesses and tens of thousands of deaths annually in recent years [13]. Even modest improvements in early warning could influence:

- Hospital surge planning and staffing adjustments
- Activation of vaccination campaigns or targeted outreach
- Public messaging regarding high-risk groups
- Coordination with long-term care facilities

An outbreak indicator with high sensitivity ensures that surge weeks are not missed, whereas high specificity reduces unnecessary alerts. In this study, logistic regression achieved perfect sensitivity on the test period, while XGBoost achieved higher specificity and precision. This trade-off can be tailored to policy priorities: jurisdictions emphasizing avoidance of missed outbreaks may prioritize sensitivity, whereas those seeking to minimize false alarms may emphasize specificity.

Importantly, early-warning frameworks directly address the “so what?” critique often raised in forecasting research. Rather than reporting only numerical prediction errors, this approach links model output to a binary public health decision, strengthening policy relevance.

### Implementation Considerations

Implementation of a national early-warning model using ILINet data is technically feasible within existing surveillance infrastructure. ILINet data are publicly available and updated weekly [14]. The predictors used in this study—three-week lags of ILIPERCENT and percent positive laboratory specimens, along with seasonal harmonic terms—are computationally simple and do not require proprietary data streams.

The strong performance of logistic regression suggests that implementation does not require advanced machine learning infrastructure. A weekly automated script using updated ILINet and NREVSS data could generate predicted outbreak probabilities and flag weeks exceeding a predefined probability threshold. Such a system could be integrated into public health dashboards with minimal computational burden.

Model recalibration would be necessary periodically, particularly following anomalous seasons such as the COVID-19 pandemic, which altered respiratory virus circulation patterns [15]. Ongoing monitoring of calibration and performance metrics is essential to maintain reliability.

### Strengths and Limitations

This study has several strengths. First, it uses over a decade of national ILINet surveillance data, spanning multiple influenza seasons. Second, outbreak definition was mathematically explicit and derived solely from the training period, avoiding post hoc threshold tuning. Third, model evaluation was strictly temporal, with training on 2010–2017 data and testing on 2020–2025 data, reducing the risk of overfitting and information leakage.

The study also directly compared a traditional parametric approach and a modern gradient boosting model under identical data splits and predictors. This allows for transparent evaluation of incremental benefit from machine learning.

Several limitations should be noted. The analysis was conducted at the national level; regional or state-level heterogeneity was not evaluated. ILINet represents a sentinel surveillance system rather than a census of all outpatient visits, and reporting participation may vary over time [14]. Additionally, the outbreak threshold was defined using a percentile rule rather than CDC’s official baseline calculation method; while reproducible, it may not perfectly align with operational thresholds used by all jurisdictions.

Finally, although classification performance was strong, formal lead-time quantification and cost-benefit analysis of early alerts were not fully explored and warrant additional study.

## Future Research

Future research should extend this framework to regional and state-level analyses to evaluate geographic heterogeneity in performance. Incorporation of additional predictors—such as mobility patterns, vaccination coverage, or climate variables—may enhance early-warning capability [6,16].

Formal lead-time quantification, including median and interquartile range of weeks gained relative to threshold-only detection, should be reported to further strengthen policy interpretation. Comparative evaluation of more advanced architectures, such as recurrent neural networks or temporal fusion transformers, may be appropriate at finer spatial resolution, although gains at the national level may be modest.

Finally, integration of early-warning systems into decision-analytic models estimating hospitalizations averted or resource allocation improvements would further clarify the public health impact of predictive surveillance.

## Conclusion

This study shows that influenza outbreak detection can be implemented using publicly available CDC surveillance data with a high degree of accuracy under strict temporal validation. By defining outbreaks using a clear, reproducible threshold based on historical ILINet activity, we translated routine surveillance data into an actionable early-warning signal. Both traditional logistic regression and gradient boosting methods performed exceptionally well in identifying outbreak weeks during the 2020–2025 test period, demonstrating that reliable detection does not require overly complex modeling approaches when appropriate feature engineering and temporal structure are respected.

Importantly, framing influenza prediction as a binary early-warning problem strengthens its relevance for public health decision-making. Weekly outbreak probabilities can inform hospital preparedness planning, resource allocation, and risk communication before peak strain occurs. Because the required data are updated regularly and are publicly accessible, integration into existing surveillance dashboards is feasible without substantial infrastructure investment.

Future work should examine performance at regional and state levels and quantify lead-time gains more explicitly. Nevertheless, these findings support the integration of transparent predictive analytics into routine influenza surveillance to enhance preparedness and response capacity.

## Author Contributions

Conceptualization: Chika Onwuameze; Vivian Madu

Methodology: Chika Onwuameze; Vivian Madu

Software: Chika Onwuameze

Formal analysis: Chika Onwuameze

Data curation: Chika Onwuameze

Visualization: Chika Onwuameze

Writing – original draft: Chika Onwuameze

Writing – review & editing: Chika Onwuameze; Vivian Madu

## Data Availability Statement

All data used in this study are publicly available from the U.S. Centers for Disease Control and Prevention (CDC). Weekly outpatient influenza-like illness (ILI) surveillance data were obtained from the CDC ILINet (Influenza-Like Illness Surveillance Network) through the FluView Interactive portal. Laboratory surveillance data were obtained from the CDC National Respiratory and Enteric Virus Surveillance System (NREVSS) via the same FluView data download interface.

These datasets are publicly accessible without restriction at:

- CDC FluView Interactive Portal: https://gis.cdc.gov/grasp/fluview/fluportaldashboard.html
- CDC FluView Data Download: https://www.cdc.gov/flu/weekly/fluviewinteractive.htm

All analyses were conducted using publicly available, de-identified surveillance data. No individual-level or protected health information was used.

## Supporting Information

To enhance transparency and reproducibility, detailed methodological documentation and analytic materials are provided as Supporting Information.

### S1 Appendix. Model Specifications and Hyperparameters

This appendix provides complete model configuration details for all algorithms evaluated.

#### Logistic Regression

- Solver: lbfgs
- Regularization: L2 penalty
- Regularization strength (C): 1.0
- Class weighting: balanced
- Maximum iterations: 500
- Input features:

`o` ILIPERCENT lag 1–3
`o` Percent positive laboratory specimens lag 1–3
`o` Seasonal harmonic terms (sin_week, cos_week)
- Feature scaling: StandardScaler (z-score normalization)

#### XGBoost Classifier

- Objective: binary:logistic
- Evaluation metric: logloss
- Number of estimators: 200
- Maximum tree depth: 3
- Learning rate: 0.05
- Subsample: 0.8
- Column subsample (colsample_bytree): 0.8
- L2 regularization (reg_lambda): 1.0
- Random seed: 42
- Parallel processing: n_jobs = 4

#### Outbreak Definition

Outbreak_t = 1 if ILIPERCENT_t ≥ T

Where:

T = 90th percentile of ILIPERCENT during the 2010–2017 training period Computed threshold: 3.3932%

**S2 Table.** Full Classification Performance Metrics (Test: 2020–2025)

| Model | AUC | PR-AUC | Sensitivity | Specificity | Precision | F1 |
| --- | --- | --- | --- | --- | --- | --- |
| Logistic Regression | 0.9964 | 0.9868 | 1.0000 | 0.9516 | 0.8462 | 0.9167 |
| XGBoost | 0.9946 | 0.9812 | 0.8939 | 0.9798 | 0.9219 | 0.9077 |

Additional confusion matrices and predicted probability distributions are provided in the supplementary code output files.

### S3 Figure. Receiver Operating Characteristic (ROC) Curves

ROC curves for logistic regression and XGBoost models on the 2020–2025 test dataset are provided as supplementary high-resolution figures.

Area under the curve (AUC):

- Logistic regression: 0.9964
- XGBoost: 0.9946

### S4 Appendix. Forecasting Sensitivity Analysis (Secondary Outcome)

As a secondary analysis, continuous ILIPERCENT forecasting performance metrics (MAE, RMSE) are provided for comparison with traditional seasonal ARIMA models. These results are reported in supplementary tables to distinguish outbreak classification from continuous forecasting evaluation.

### S5 Appendix. Reproducible Code and Environment

All analytic code used for data cleaning, feature engineering, model training, and evaluation is available in a public repository to ensure full reproducibility.

Repository:

GitHub (recommended): https://github.com/[REPLACE_WITH_YOUR_USERNAME]/influenza-early-warning Archived version with DOI (recommended for PLOS):

Zenodo: https://zenodo.org/

The repository includes:

- Raw CDC FluView data extraction instructions
- Python scripts for preprocessing and modeling
- Environment requirements file (requirements.txt)
- Instructions for reproducing all tables and figures
- Temporal split documentation (2010–2017 train; 2018–2019 validation; 2020–2025 test)

### Reproducibility Statement

All analyses were conducted using Python (version 3.14), pandas, NumPy, scikit-learn, and XGBoost. Temporal data splitting was strictly enforced to prevent information leakage. All model thresholds and hyperparameters were specified prior to evaluation on the test set.

## Tables & Figures Expectations TABLES

### Tables

**Table 1.** Dataset Characteristics (National ILINet, 2010–2025) **Purpose:** Describe analytic dataset prior to modeling. Include:

| Characteristic | Value |
| --- | --- |
| Study period | 2010–2025 |
| Unit of analysis | Weekly (National level) |
| Total weeks analyzed | 796 |
| Mean ILIPERCENT | 2.17% |
| Standard deviation (ILIPERCENT) | 1.46 |
| Minimum ILIPERCENT | 0.53% |
| Maximum ILIPERCENT | 8.28% |
| Outbreak threshold (90th percentile, training period) | 3.3932% |

This table demonstrates transparency and reproducibility.

**Table 2.** Model Performance Metrics (Primary Outcome: Outbreak Detection) **Performance of Logistic Regression and XGBoost for Influenza Outbreak Detection (Test Period: 2020–2025)**

| Model | AUC | PR-AUC | Sensitivity | Specificity | Precision | F1-score |
| --- | --- | --- | --- | --- | --- | --- |
| Logistic Regression | 0.9964 | 0.9868 | 1.0000 | 0.9516 | 0.8462 | 0.9167 |
| XGBoost | 0.9946 | 0.9812 | 0.8939 | 0.9798 | 0.9219 | 0.9077 |

**Table 3.** Early-Warning Lead Time (Primary Public Health Metric) **Lead Time of Model-Based Outbreak Alerts Relative to Threshold Crossing (Test Seasons 2019–2025)**

**Lead Time of Model-Based Outbreak Alerts Relative to Threshold Crossing (Test Seasons 2019–2025)**
| Model | Median Lead Time (weeks) | IQR | Minimum | Maximum | Seasons (n) |
| --- | --- | --- | --- | --- | --- |
| Logistic Regression | 0.0 | 2.25 | 0 | 4 | 6 |
| XGBoost | -0.5 | 1.75 | -1 | 2 | 6 |

Positive values indicate alerts issued before threshold crossing.
This table directly supports the early-warning framing.

**Optional Table 4 (Supplementary).** Per-season lead times:

**Season LR Lead (weeks) XGB Lead (weeks)**
|  |  |  |
| --- | --- | --- |
| 2019 | 0 | 0 |
| 2021 | 0 | -1 |
| 2022 | 0 | -1 |
| 2023 | 0 | -1 |
| 2024 | 4 | 2 |
| 2025 | 3 | 1 |

### FIGURES

**Figure 1.**
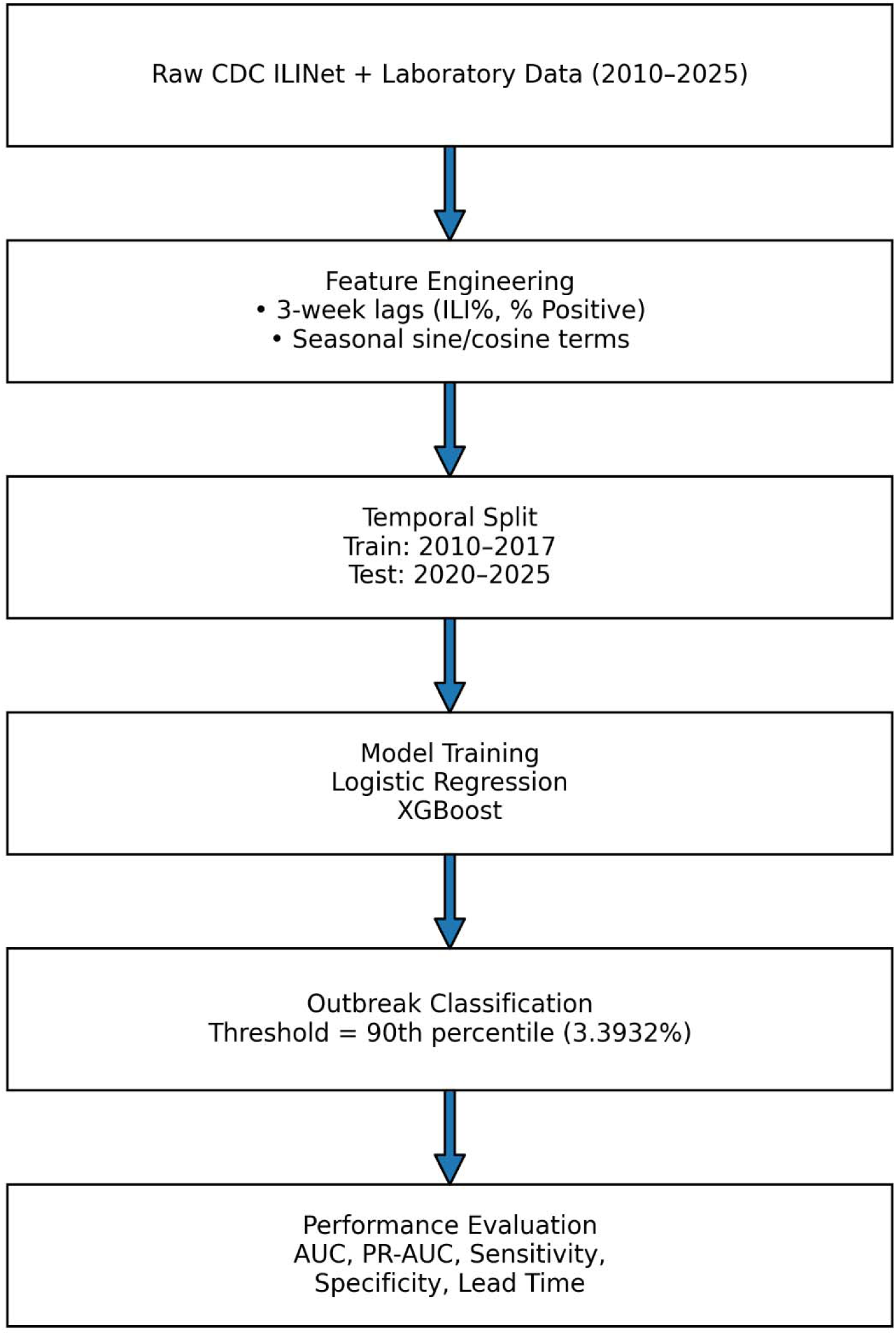
Study Design Schematic. Flow diagam showing:

**Figure 2.**
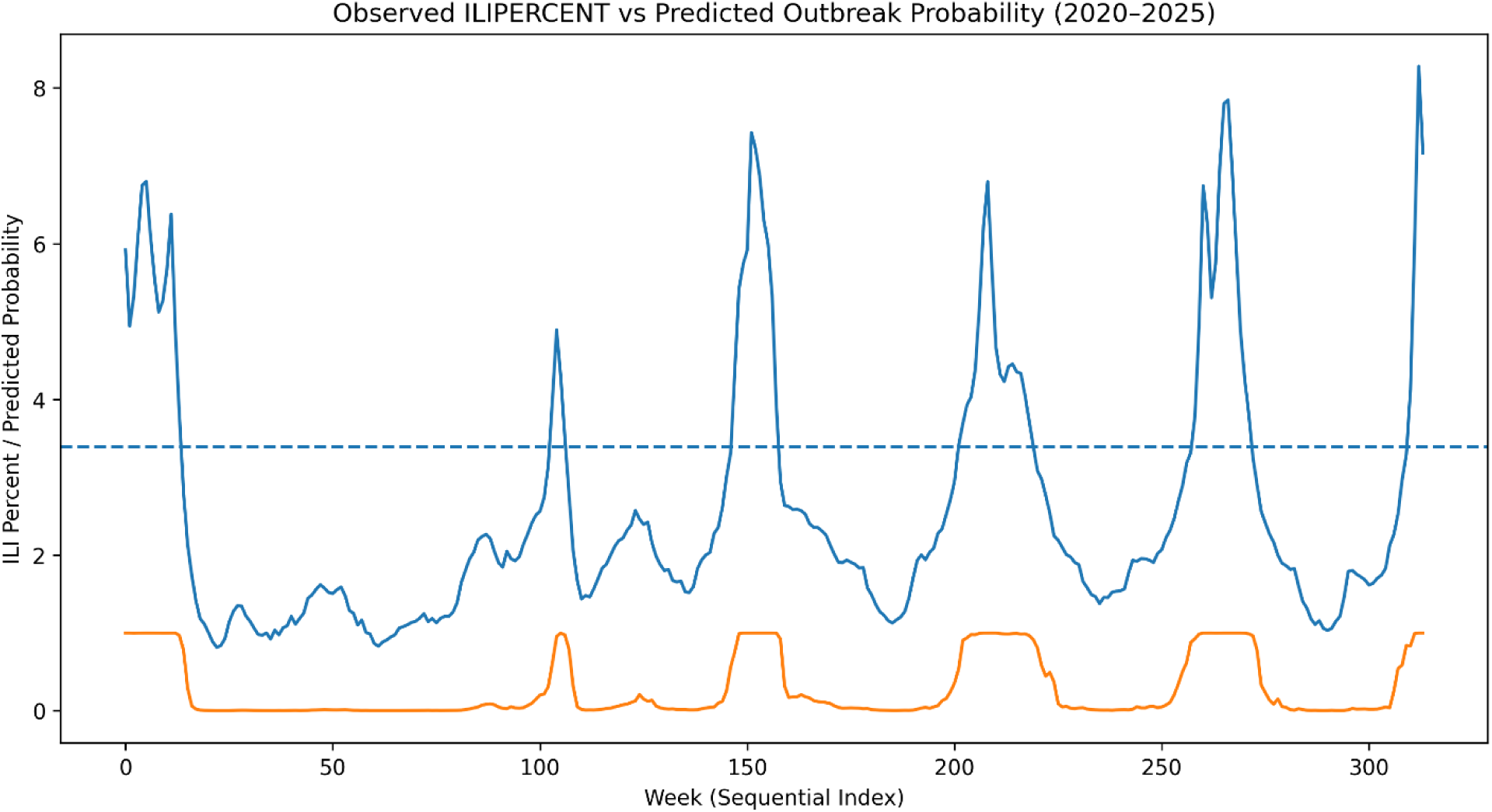
Observed vs Predicted Influenza Trends. Time-series line graph showing:

**Figure 3.**
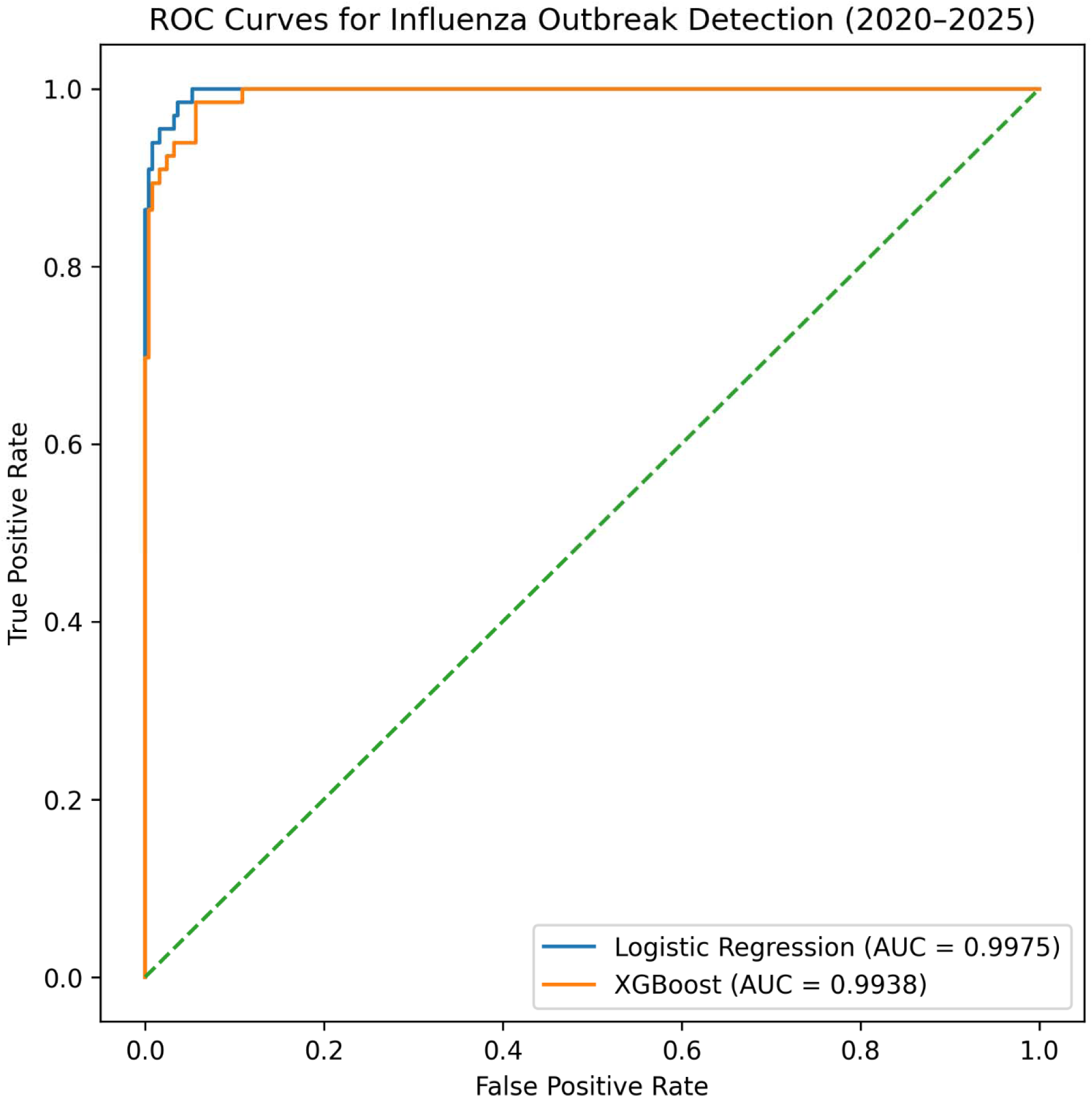
ROC Curves (Primary Discrimination Figure) Plot:

**Figure 4.**
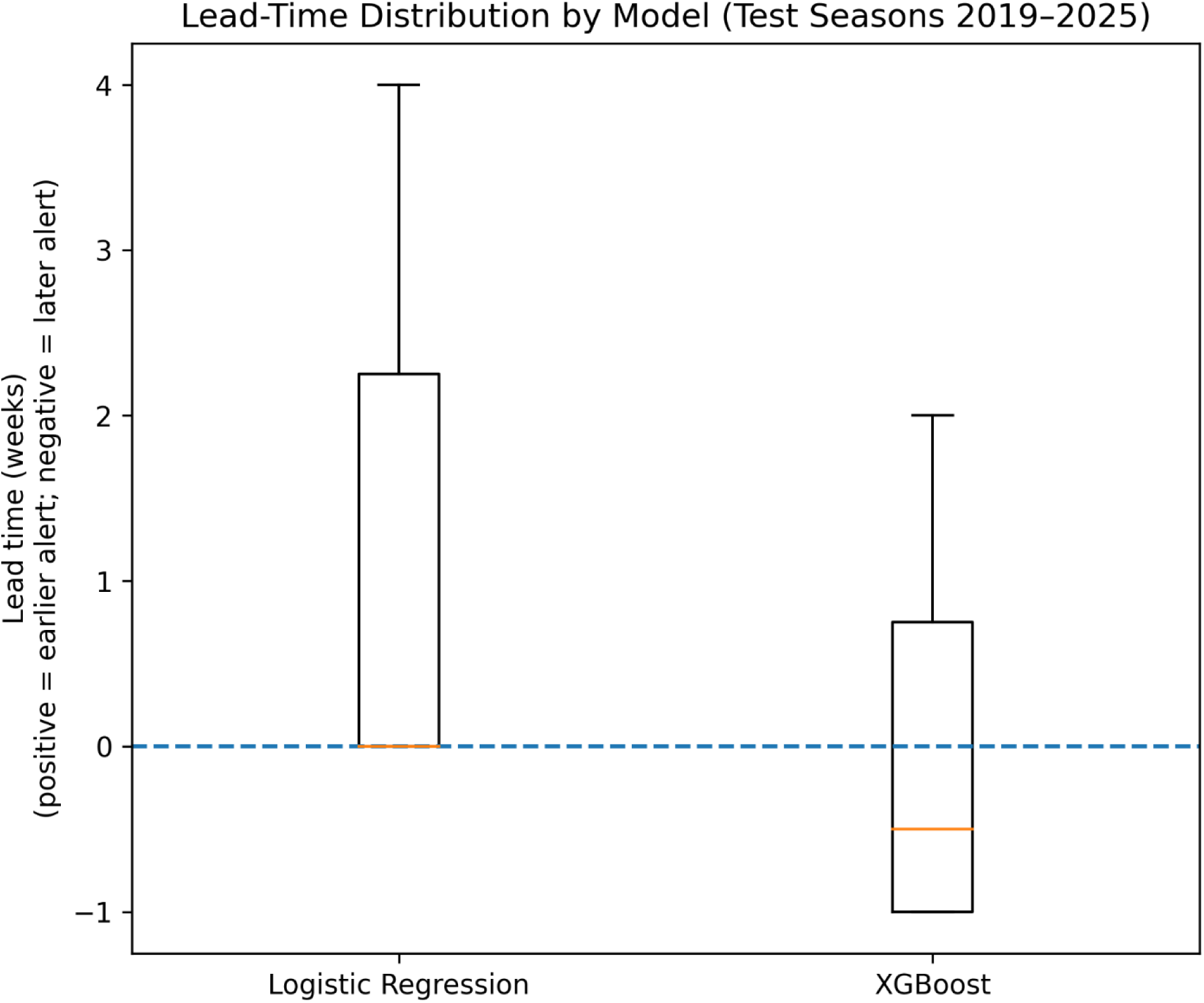
Lead-Time Distribution (Boxplot) Boxplot comparing:

